# A first study on the impact of current and future control measures on the spread of COVID-19 in Germany

**DOI:** 10.1101/2020.04.08.20056630

**Authors:** Maria Vittoria Barbarossa, Jan Fuhrmann, Julian Heidecke, Hridya Vinod Varma, Noemi Castelletti, Jan H. Meinke, Stefan Krieg, Thomas Lippert

**Affiliations:** Institute for Applied Mathematics, Heidelberg University, Heidelberg, Germany; Interdisciplinary Center for Scientific Computing, Heidelberg, Germany; Frankfurt Institute of Advanced Studies, Frankfurt, Germany; Institute of Radiation Medicine, Helmholtz Zentrum München, Neuherberg, Germany; Jülich Supercomputing Centre, Forschungszentrum Juülich, 52425 Jülich, Germany

## Abstract

The novel coronavirus (SARS-CoV-2), identified in China at the end of December 2019 and causing the disease COVID-19, has meanwhile led to outbreaks all over the globe, with about 571,700 confirmed cases and about 26,500 deaths as of March 28th, 2020. We present here the preliminary results of a mathematical study directed at informing on the possible application or lifting of control measures in Germany. The developed mathematical models allow to study the spread of COVID-19 among the population in Germany and to asses the impact of non-pharmaceutical interventions.

## 1. Introduction

The novel coronavirus (SARS-CoV-2), identified in China at the end of December 2019 and causing the disease COVID-19, has meanwhile led to outbreaks all over the globe, with about 571,700 confirmed cases and about 26,500 deaths as of March 28th, 2020 [1]. We present here the preliminary results of a mathematical study directed at projecting possible modifications and improvement of control measures in Germany. The developed mathematical models allow to study the spread of COVID-19 among the population in Germany and to asses the impact of non-pharmaceutical interventions. The overall goal is to suggest strategies for the mitigation of the current outbreak, slowing down the spread of the virus and thus reducing the peak in daily diagnosed cases, the demand for hospitalization or intensive care units admissions, and eventually fatalities. The project was developed in the framework of a newly established collaboration on “Epidemiology and Pandemics” between Heidelberg University, the Frankfurt Institute of Advanced Studies and the Jülich Supercomputing Center.

## Highlights

- In this study, mathematical modeling is used to predict the spread of COVID-19 among the German population and to evaluate the impact of non-pharmaceutical interventions. The proposed setting allows to investigate how a specific intervention scenario affects the dynamics of the epidemics, with particular attention to interactions between individuals of the same or different age groups (children, adults and people older than 60y).
- **A most undesirable scenario (do-nothing scenario)**, where the only control measures are increased awareness due to media activities (washing hands, proper coughing and sneezing, keeping distance from obviously sick persons), according to the mathematical model would lead to a peak in the curve of diagnosed cases at the end of April 2020, with about 2.8 Mio active detected cases on the day of the peak, a total of 80 Mio infected (out of which only about 8 Mio detected and 23.5 Mio asymptomatic SARS-CoV-2 infections), and a total of 630,000 fatalities over the course of the epidemic. This is in line with what was predicted for the outbreak in the United Kingdom in an analogous do-nothing scenario [4].
- **Hitherto adopted main control measures** (incl. closure of schools and universities, remote working policy, isolation of infected cases and maintenance of testing activity as of March 2020) have then been included in the mathematical model to fit the curves of reported cases in Germany as of March 25th. **The maintenance of these measures until the end of 2020 was assumed to predict the “baseline” (BSL) scenario**. Model simulations indicate that, compared to the do-nothing scenario, where no interventions are applied, the BSL scenario would lead to: (i) a shift in the epidemic peak by about one month (expected in early June 2020), (ii) a reduction of detected SARS-CoV-2 infections by 60% (from 2.8 Mio to 1.26 Mio) at the outbreak peak, (iii) a reduction by about 100,000 fatalities (expected over 530,000 fatalities in total) and (vi) about 69 Mio infected (thereof 20 Mio asymptomatic SARS-CoV-2 infections) over the course of the epidemic.
- **A low-damage scenario could be predicted by enriching the baseline measures with significantly increased testing activity** (not only suspected SARS-CoV-2 infectives but also persons without symptoms or known close contacts to identified cases), a rather strict isolation protocol of detected SARS-CoV-2 cases for about two weeks, and reduced contacts with vulnerable individuals. According to the model, these interventions, if starting immediately and going into full effect within five to ten days seem to reduce the number of fatalities to a minimum of 18,000 and the peak number of infectives to 660,000 active detected infections at the day of the peak in the third week of April. However, this dramatic slowdown of the epidemic comes with a price since successfully preventing new infections also slows down the increase of the number of recovered, and hence immune, individuals. **For this scenario to work effectively, all the above mentioned strict control measures (including BSL measures) must remain in effect for more than one year**.
- A partial lifting of the restrictions imposed thus far, planning for a **reopening of schools, universities, and restaurants, resuming work and most club activities after mid April 2020** is predicted to result in an increase of some 15% of the death toll over the BSL scenario by the model. Simulations indicate that the favorable effects of the restrictions imposed to date would be substantially reversed, leading to a comparable scenario as the do-nothing scenario, with an epidemic peak around mid May 2020, 2.2 Mio diagnosed cases on the day of the peak and 620,000 fatalities over the course of the epidemic. **If, however, this is accompanied by significantly increased testing activity, isolation of identified cases and reduced contact with vulnerable individuals, such a partial withdrawal leads to less severe consequences**. Indeed, model simulations of this scenario indicate that the (first) epidemic peak would be reached due to increased testing activity in the second half of April 2020, with 670,000 detected SARS-CoV-2 infections at the outbreak peak. A second peak would follow and the epidemic would last for a longer period (more than one year), but the total number of cases (14.4 Mio, out of which there are 1.2 Mio asymptomatic SARS-CoV-2 infections) and fatalities (about 60,000) would be substantially limited by the measures according to the model.
- **A close-to-total shutdown of any economic and social activities for a period of 5 weeks from now** will probably only cause a shift in one of the described scenario. While the advantage of a temporary shutdown is buying time (the expected peak being mid July 2020 with 1.3 Mio identified cases at the day of the peak), which might be helpful if the health care systems can be prepared to be in a better position to deal with the disease, such a scenario would still lead to about 570.000 fatalities in total, as the model predicts. If accompanied by increased test activity this scenario leads to similar fatality numbers (258,000) and even higher peak numbers of known infected individuals (22.6 Mio) as compared to a scenario with increased testing alone as additional measure. The main effect of a shutdown with abrupt start and end would be delaying the peak of known active cases, while at the same time making it narrower and higher. **A significant increase in testing activity for the detection of infections, including for antibodies against SARS-CoV-2 could be quite beneficial**. First, this will reveal a large number of already present but yet undetected infected individuals, and allow to carefully isolate them. Further, antibody testing would allow identifying unknown recovered (presumably immune) individuals, who could then resume their normal levels of activity.

## 2. Methods

### 2.1 Data

The meanwhile publicly available dataset provided by the Robert Koch Institute (RKI) [2] was used for this study and was last accessed on March 26th, 2020. Total cases per federal state and district are recorded in the database as reported to the local health authorities. The reporting date - the date on which the local health authority became aware of the case and recorded it electronically - is used to display the newly transmitted cases per day. A few days can pass between the notification by the doctors and laboratories to the health authority and the transmission of the data to the RKI, henceforth the exact time of infection of the reported cases cannot usually be determined. The notification date to the health department therefore best reflects the time of detection of the infection (diagnosis date) and thus the current infection process.

The dataset is provided as a grouped incidence table where each stratum (data row) is defined by the variables: “Federal State” (Bundesland), “District” (Kreis), “Age” (categorical), “Gender” and “Reporting date”. Six different age groups are reported: 0-4 years, 5-14 years, 15-34 years, 35-59 years, 60-79 years and 80+ years. For 0.4% of cases the age information is missing. The cumulative number of reported SARS-CoV-2 infections in Germany up to March 26th was confirmed to be 36,508, with +4,953 (+13.6%) new cases compared to the previous day. The percentage of male patients (54.3%) is higher compared to that of females (45.3%), (0.4% gender unknown). A similar feature was observed also in COVID-19 data from other countries, e.g. Italy [3]. Most patients are registered with an age between 15 and 79 years, with the predominance of the age category 35-59 (Figure 1a). The earliest cases were reported on Jan 24th, 2020. The first dead patients were reported on Feb 20th, 2020 (Figure 1b). Since March 4th, 2020 (with the exception of March 8th, 2020) at least one death per day was reported. The age distribution of dead patients is dominated by the age category 80+ years. As of March 26th no fatal case younger than 35 years was reported.

**Figure 1:**
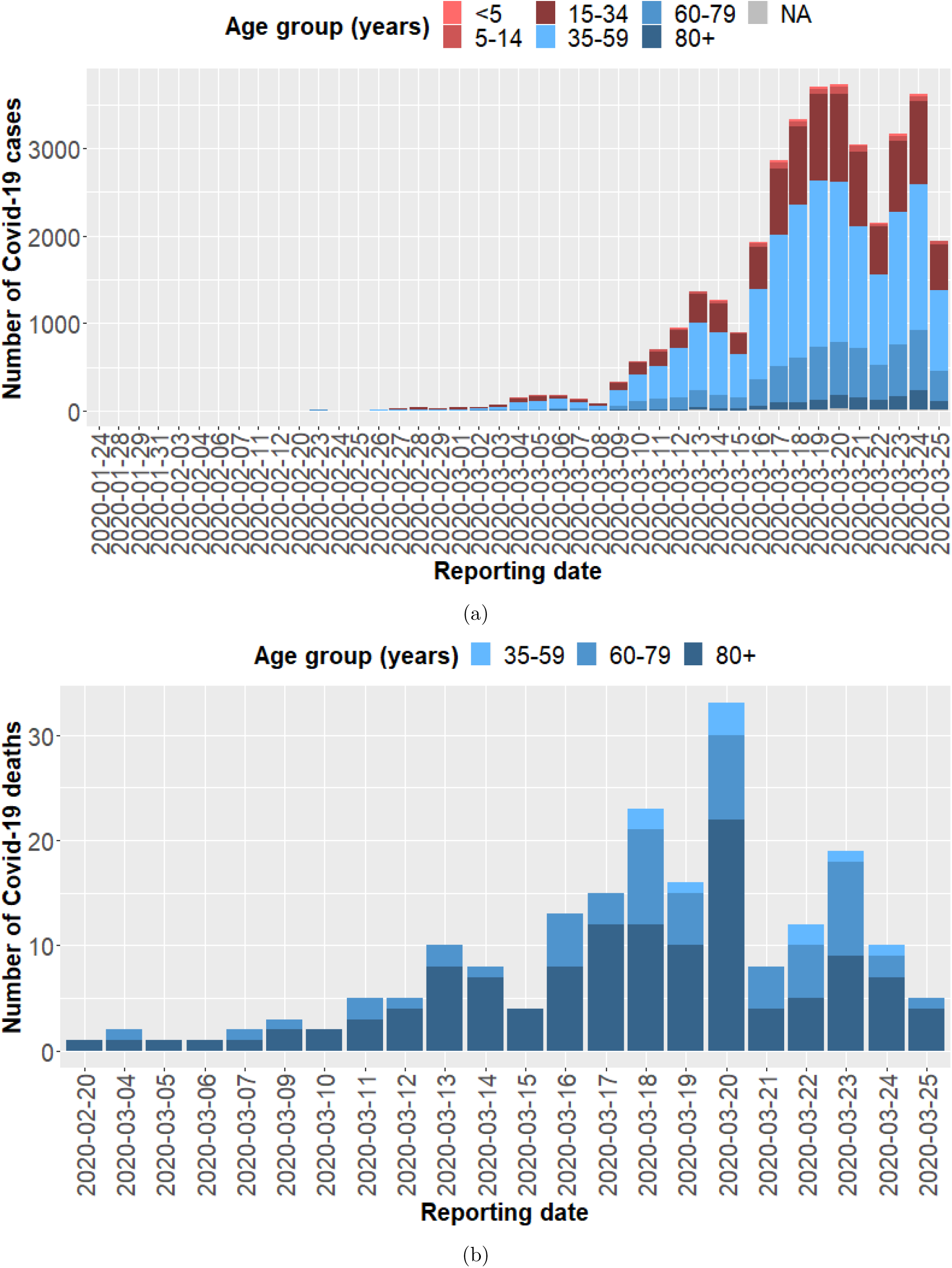
Data from [2] as of March 26th, 2020. (a) Daily new COVID-19 cases classified by age groups reported in Germany since Jan 24th, 2020. (b) Daily new COVID-19 deaths classified by age groups reported in Germany since Feb 20th, 2020.

### 2.2 Mathematical Models

The mathematical models adopted for this study are based on systems of differential equations which describe interactions between different groups of individuals in the population. The proposed approach extends the known S-E-I-R (susceptibles-exposed-infected-recovered) scenario for disease dynamics [7]. Individuals are classified according to their status with respect to the virus spread in the community. In particular we distinguish between individuals who have been exposed to the virus but are not yet infectious, asymptomatic infectives, infectives with mild or influenza-like symptoms (not reported as SARS-CoV-2 infections), and reported SARS-CoV-2 infectives. We assume that infected patients without SARS-CoV-2 diagnosis are unlikely to die of the virus-induced disease. Undetected infections lead to undetected recoveries in the population, which cannot be reported unless testing on ongoing (virus detection) or previous (antibody detection) SARS-CoV-2 infections is performed.

The so-called basic reproduction number (ℛ_0_) of an infectious disease is a parameter which can be calculated from the underlying mathematical model of the transmission network. In simple disease transmission models ℛ_0_ *>* 1 implies that the virus will spread in the population, and the larger the value of ℛ_0_, the harder it is to control the epidemic.

### 2.3 Preliminary study - Elementary model with homogeneous population

The simplest approach that we adopted to understand the evolution of the epidemic in time is based on the assumption that the population is homogeneous (in particular with respect to age and social habits). This simplistic assumption is of course not reflecting the multidimensional complexity of the ongoing situation but can nevertheless help in determining major factors affecting disease spread. Calibrating our simplest (homogeneous) model on time series for reported cases (data from [2]) from January 28th until March 13th, 2020 we obtain ℛ_0_ = 4.3. This value is obtained assuming minimal or no control measures applied to mitigate the spread of the epidemic and is in line with previous estimates for ℛ_0_ in other countries during the initial phase of the outbreak [5, 6]. Figure 2 shows the situation without any control measure applied, as the epidemic spreads among the population. The severity of the outbreak is highly dependent on contacts with infectives (symptomatic or not). A reduction in contact rates of all individuals postpones and lowers the peak in the curve of currently active infections (cf. Fig. 3(a)). Under our working hypotheses, asymptomatic individuals are those with highest contact rates (due to absence of symptoms), while detected COVID-19 cases are those with lowest contact rates, though not being completely isolated. Reduction of contacts of asymptomatic infectives, as well as successful measures for early detection of new infections are potential measures to reduce the basic reproduction number, hence the severity of the outbreak (cf. Fig. 3(b)).

**Figure 2:**
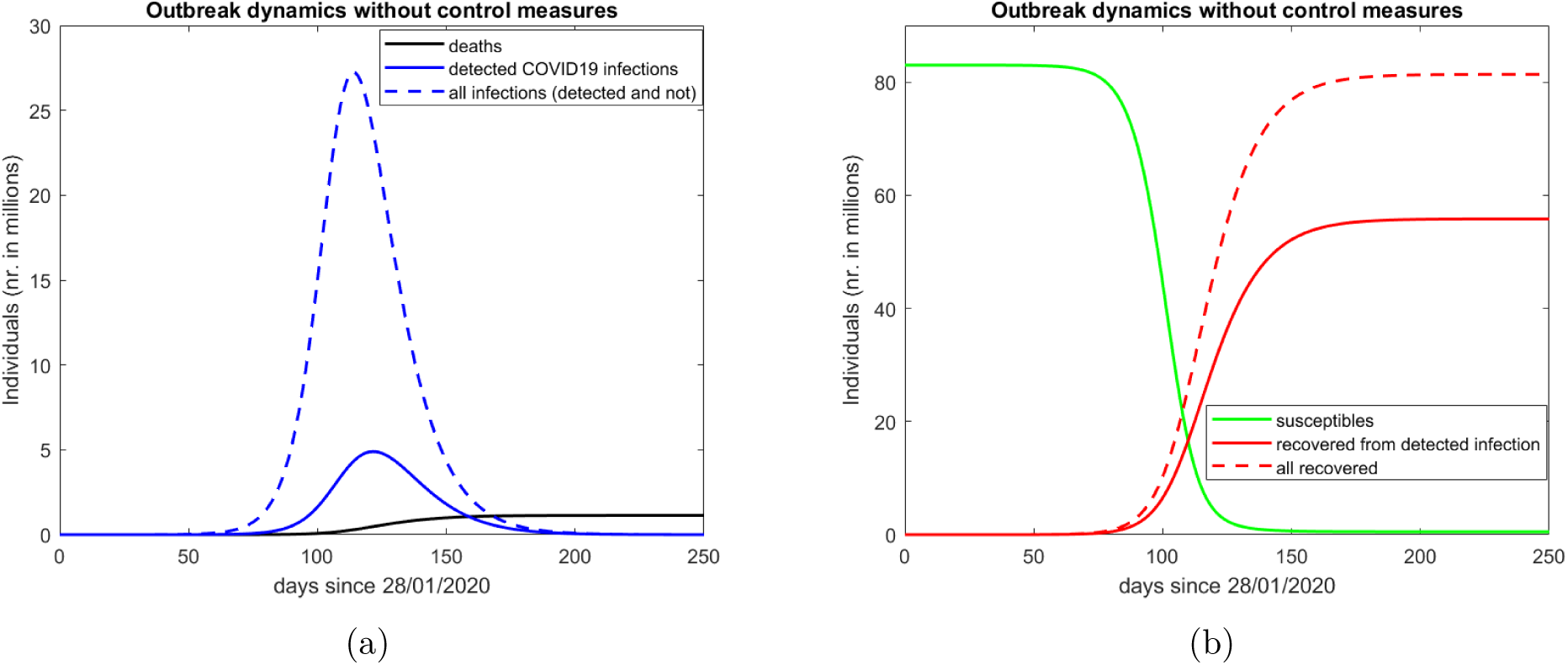
COVID-19 outbreak scenario in absence of active control on virus spread. Elementary model with parameters calibrated to German data from [2] collected until March 13th, 2020. This model does not take into account age or social structures in the population and is used here only as a proof of concepts for the refined modeling approach.

**Figure 3:**
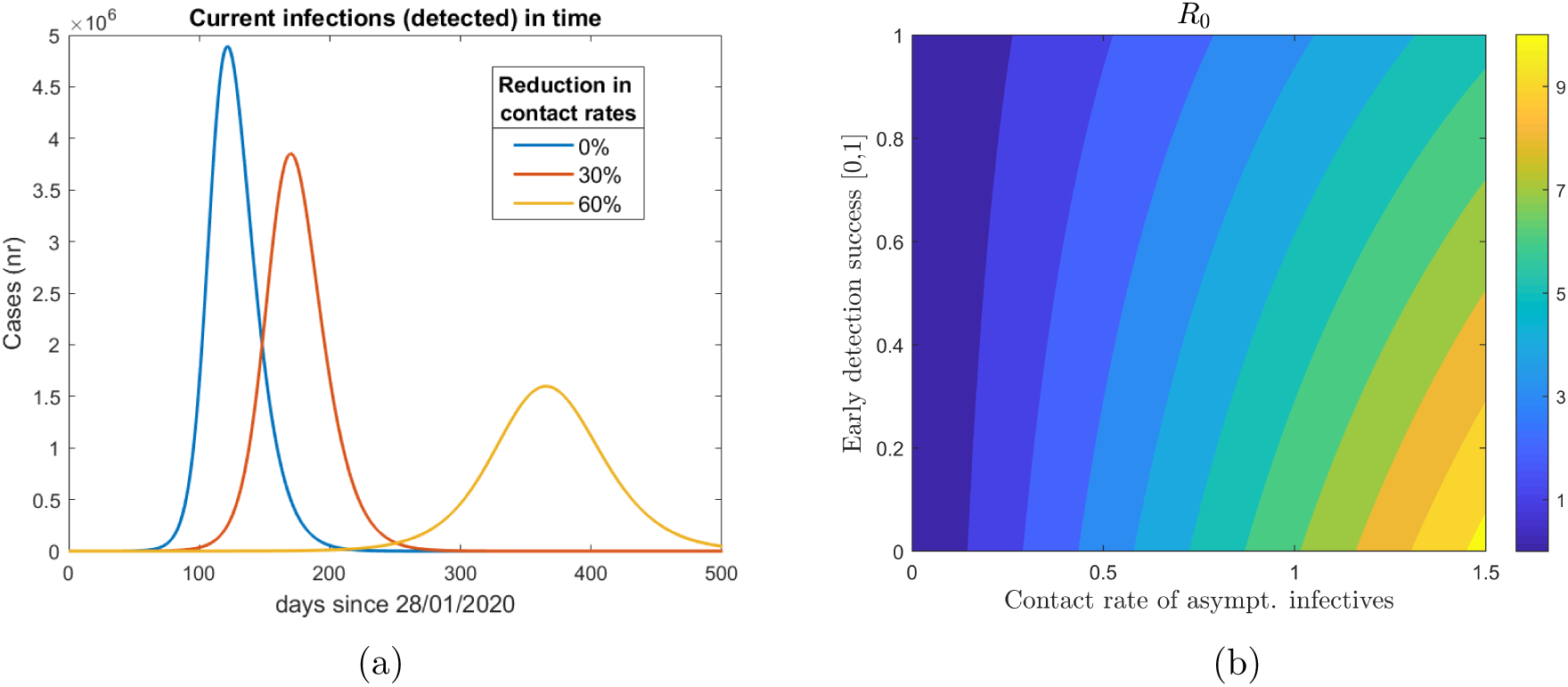
Reduction in contact rates, in particular of asymptomatic infectives, and early detection of new infections (a) lead to a less severe outbreak and (b) can be used to control the basic reproduction number ℛ_0_.

### 2.4 Refined model

The preliminary model described in Sec. 2.3 was refined to include age groups and immunological stages during infections. This allows to consider features like the immune response of individuals during infection, as well as social behavior, in particular interactions among individuals of the same or different age groups. Based on the statistical analysis of the RKI data 2.1 we distinguish three groups: children (0-14y), adults (15-59 y) and people 60y or older. The refined model also includes additional stages of infection to obtain a more realistic time course of individual infections.

Further we incorporate in the model (i) general increased awareness in the population due to the effect of media (M), as well as (ii) active control due to main intervention measures adopted in Germany since February 2020. These control measures include: (CS) Closure of all schools, universities, sports clubs and canceling public events; (HO) reduced contacts in workplaces and outside the household (restaurants, bars, public transport); (T0) initial efforts to improve detection by more testing, and (IC0) isolation of infected cases. Details are summarized in Table 1. Collected reported cases for the three age groups were used to fit the model including control measures as indicated in Table 1, until March 25th (Fig. 4a). The obtained setting was used to construct predictions (Fig. 4b) the baseline (BSL) scenario, in which the currently applied control measures (CS,HS,TO, and ICO) and the awareness due to the effect of media are maintained for the time of simulations (1 year). **The BSL scenario is projected to result in more than half a million fatalities (648 deaths per 100**,**000 popul.) throughout Germany, with over 1.2 mio active cases on the day of the peak (day 127, beginning of June 2020)**. Compared to a scenario without the application of interventions, the maintenance of the baseline measures until the end of 2020 leads to a reduction in about 100.000 fatalities and a reduction of number of detection at the outbreak peak by 60% (from 2.8 Mio to 1.26 Mio.).

**Table 1:** Summary of control measures for mitigation of COVID-19 infections adopted in Germany as of March 26th.

| Label | Policy | Description/Working assumptions |
| --- | --- | --- |
| CS | Closure of all schools, universities, sport clubs. Public events are canceled. | Reducing contacts: child-child (- 60%), child-adult (-5%), adult-adult (- 50%), senior-senior (- 10%). Assumed to be applied at national scale on March 14th 2020 (day 48). |
| HO | Remote working policy (home office), closure of all restaurants and bars, reduced (use of) public transport service. | Reducing contacts: child-child (- 20%), child-adult (- 20%), adult-adult (- 50%), child-senior (- 30%), adult-senior (- -30%), senior-senior (- 30%). Assumed to be applied at national scale from March 13th (day 45), contact reductions fully achieved after 13 days. |
| M | Awareness raising due to media, canceling big events | Information and media activities increase social distancing and personal hygiene (e.g., washing hands), limited (self-)quarantine of known or suspected cases starting on Feb 25th (day 28). All contacts are effectively reduced by 20%, contact between senior and adult/child slightly more reduced, fully achieved after 20 days. |
| T0 | Testing | Increased testing activity since Feb 28th (day 31). Approx. 10-fold increase in testing compared to previous phase, achieved after 21 days. |
| IC0 | Isolation of infected cases | Identified infected cases isolated for 2 weeks. Reduces contacts (for individuals in specific groups by - 70%) to be implemented starting on March 28th (day 60), fully in effect by day 65. |

**Figure 4:**
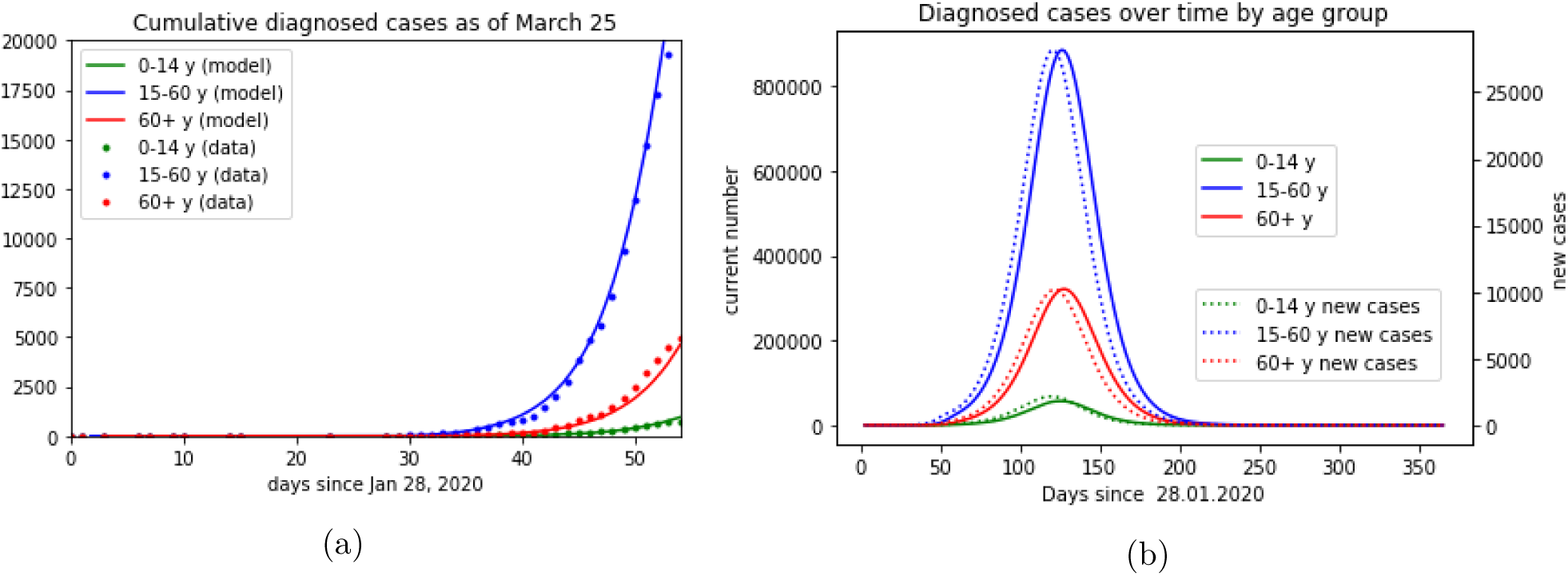
Cumulated COVID-19 cases classified by age groups. (a) Model simulations (continuous curves) and data from [2] as of March 25th, 2020. The most recent data points are highly affected by uncertainties (underreporting) and are not taken into account for model calibration; (b) Model predictions for baseline (BSL) scenario until end Jan. 2021. Dashed curves represent daily detected new cases, continuous curves represent active infections.

In light of these results and considering that the current restrictions on public life are hard to sustain in the long run, we are led to simulate several scenarios based on possible further nonmedical interventions (cf. Table 2). Among these interventions are massive expansions of testing for infections both among symptomatic (T1) and increasingly among asymptomatic persons (T1+), even stricter isolation of identified cases (IC, IC+), increased caution in contact by and with endangered individuals to different extents (IO, IO-), and finally a close to total shutdown (SD) of economic activities, as it was successfully exercised in Hubei province in China. Moreover, **we investigate the effects of at least a partial rollback of the measures being currently imposed**, specifically reopening schools, universities and some organized social activities (RB-CS) on the one hand, and vastly reduced restrictions of economic activities (RB-HO) on the other hand.

**Table 2:** Possible measures included in simulated scenarios, starting from March 28th or later.

| Label | Policy | Description/Working assumptions |
| --- | --- | --- |
| T1 | Significant increase of testing, in particular of individuals with relevant symptoms and/or contact to identified cases | Further increase in testing by at least four-fold, starting on day 60 (28.03.), fully achieved after 10 days. |
| T1+ | Significant increase in testing, *including of asymptomatic individuals without known direct contact to identified cases* | Further increase of tests by a factor of 15 (over T0), starting at day 60 (March 28th), fully implemented by day 70 (April 7th). |
| RB | (partial) Rollback of measures HO and/or CS, significantly weakens the control measure described therein | Turns back 80-90% of the reductions in contact rates achieved by HO/CS, slowly starting on day 75 (April 12th), fully in effect on day 82 (April 19th). |
| IC | Stricter isolation of known infectives compared to IC0 | Reduces contact rates of identified COVID-19 infectives by -70% (compared to IC0), starting on day 60 (March 28th), in full effect by day 65 (April 2nd). |
| IC+ | Even stricter isolation of known cases | Analogous to IC, leading to higher reduction (-90% compared to IC0) in contact rates of identified COVID-19 infectives (-90% compared to IC0). |
| IO | General social distancing (incl. self-separation) of individuals at risk, in particular elderly | Reduction of child-senior and adult-senior contacts by -70%, starting on day 60, in full effect on day 65. |
| IO- | Lighter version of IO | Reduction of child-senior and adult-senior contacts by -40%, starting on day 60, in full effect on day 65. |
| SD | Temporary close-to-total shut-down of economic and social activities | Further reduction of contacts by 80% starting immediately and being in effect for 5 weeks |

## 3 Results and Observations

Among the simulated scenarios (Fig. 5), the most coherent predictions are the effectiveness of enhanced testing (T1, T1+) and particular care toward endangered persons (IO, IO-) for the reduction of the predicted number of fatalities. The expansion of testing capacities by a factor of 10 alone already results in a reduction of fatalities by about 50% as compared to the baseline (result not shown in figures). Clearly, increased testing will result in a higher number of diagnosed cases, increasing the peak number of known infectives by a factor of two (Fig. 6b, Fig. 6c), but it is important to keep in mind that the additional identified cases should be expected to be mostly weakly symptomatic. The expected peak number of patients requiring hospitalization is reduced by more than 30%, and the same is true for patients requiring intensive care (result not shown in figures).

**Figure 5:**
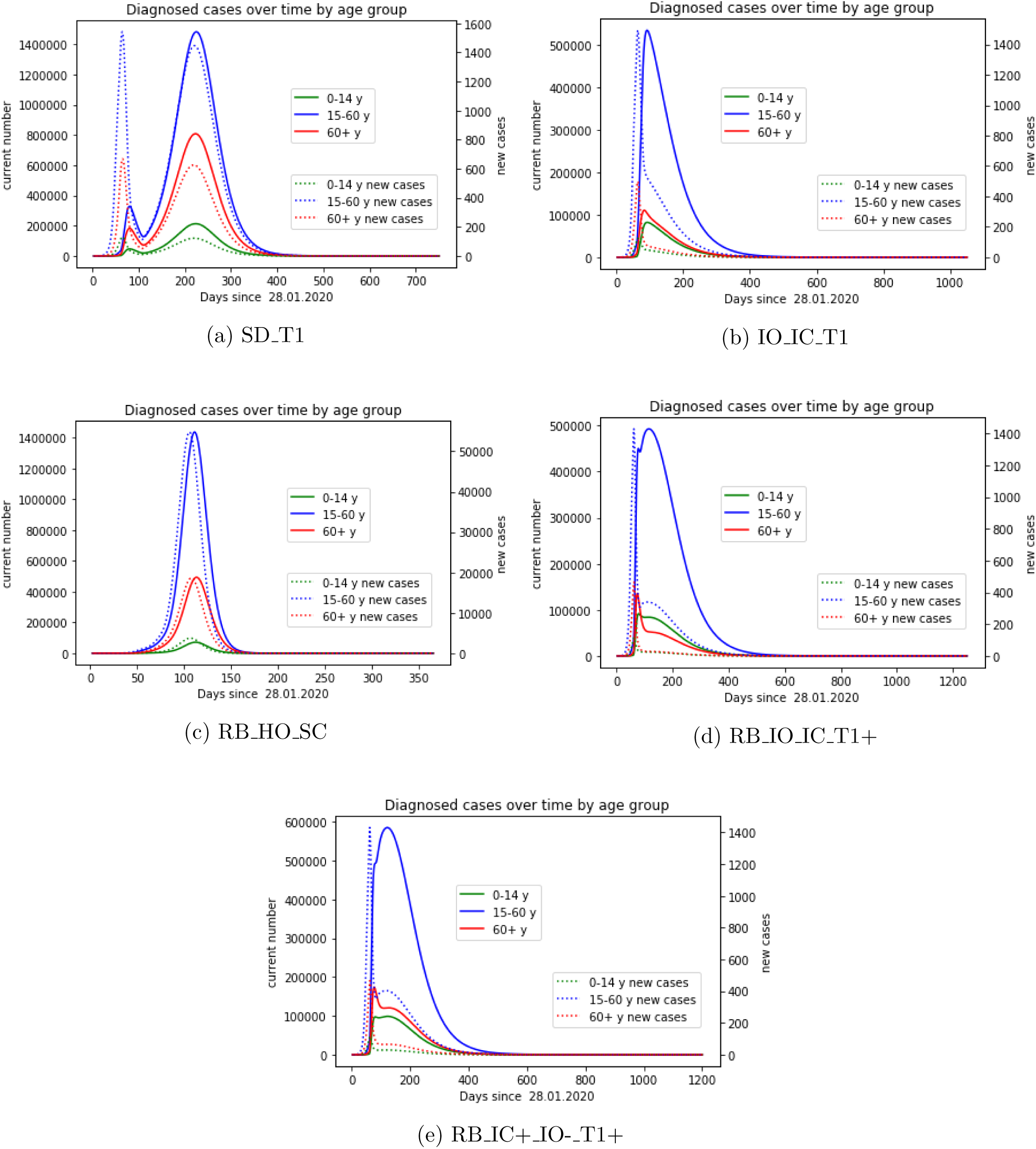
Model prediction for different model scenarios. (a) Temporary shut-down and increase testing activity (SD T1); (b) Restricted contacts with elderly and risk groups, isolation of detected cases, combined with increased testing activity (IO IC T1); (c) Partial rollback, reopening schools, reintroducing contacts at workplaces and public transportation (RB HO SC); (d) Partial rollback as in (c) coupled to strongly increased testing also of asymptomatic individuals, isolation of identified cases, and reduced contacts with elderly and risk groups (e) Partial rollback as in (c) coupled to strongly increased testing also of asymptomatic individuals, strict isolation of identified cases, and partially reduced contacts with elderly and risk groups.

**Figure 6:**
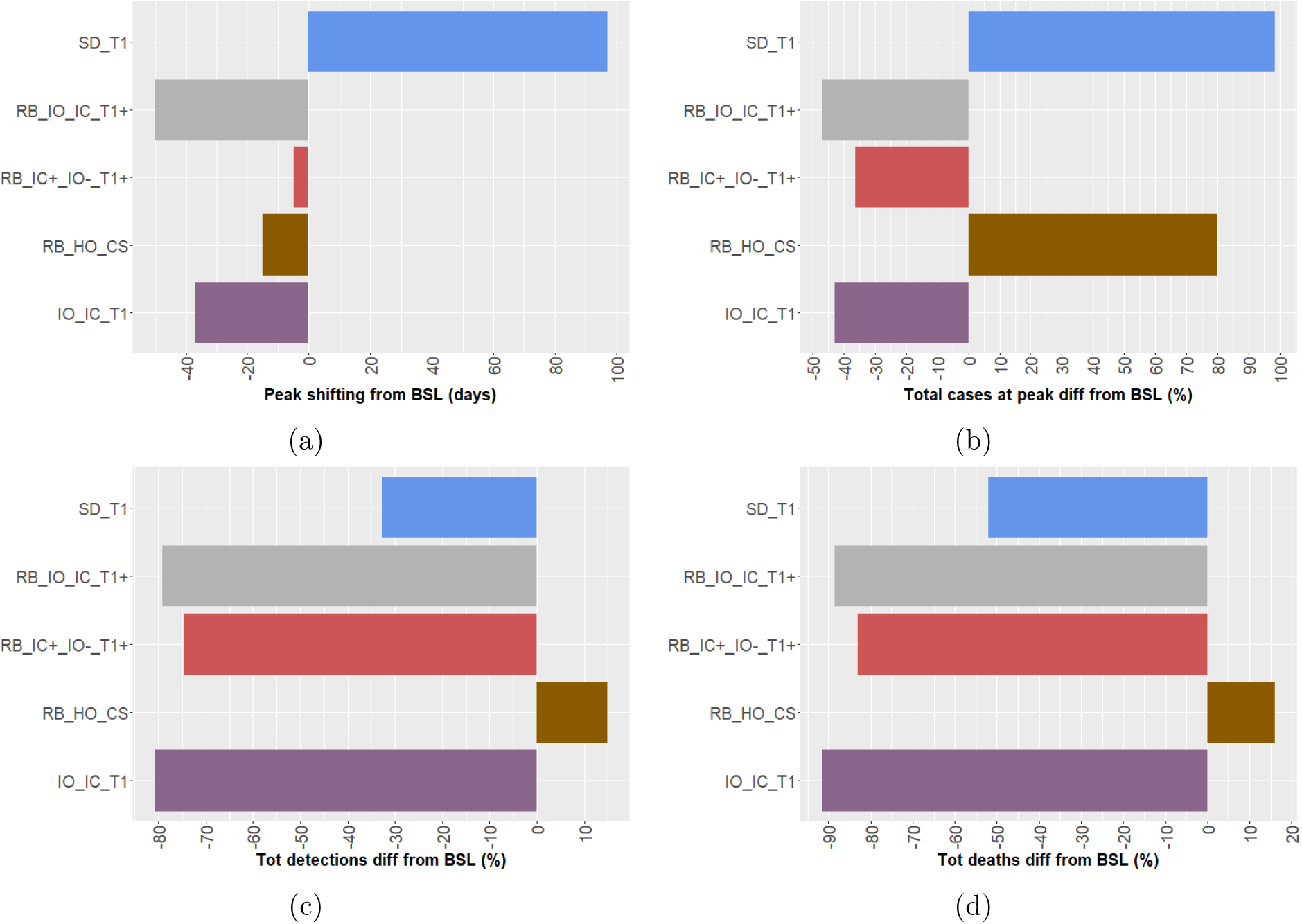
Statistical comparison of model output for the baseline (BSL) scenario and the considered possible alternatives. (a) Peak shifting (in days) compared to BSL; (b) Differences in reported cases (%) at the day of the peak; (c) Differences in total detected cases (%); Differences in total deaths (%).

**Testing not only suspected infectives but also persons without symptoms or known close contacts to identified cases is projected to reduce the number of fatalities** even further, in particular if combined with rather strict isolation of identified cases for about two weeks and strongly reduced contacts between endangered individuals and possible infectives (scenario IO IC T1+). **These interventions, if starting immediately and going into full effect within five to ten days (Table 2)**, **promise to reduce the number of fatalities by a factor of about 30 (18**,**000 expected deaths, Fig. 6c) and the peak number of infectives by a factor of two (660**,**000 active detected infections at the day of the peak Fig. 6b)**. The number of required hospitalizations and intensive care patients should be reduced even further, since the **increase in testing results in more diagnosed cases with mild or possibly without symptoms**. However, this dramatic slowdown of the epidemic comes with a price since successfully preventing new infections also slows down the increase of the number of recovered, and hence immune, individuals. It should be remarked that the current restrictions (BSL) on public life were assumed to remain largely in effect in this scenario. Save for quick breakthroughs in the development of a vaccine or effective medication, or a significant reduction of the virus’ aggressiveness, **we are talking about years rather than months during which the measures have to remain in effect**.

This leads us to consider two more scenarios which include an at least partial lifting of the restrictions imposed thus far. We assume this to start around Easter (April 12th, 2020), including reopening schools, universities, and restaurants, resuming normal work and most club activities, and at least some resurgence in travel. **Without any further intervention, this lifting of measures alone (scenario RB-HO-CS) is expected to result in an increase of some 15% of the death toll over the baseline scenario** (Fig. 6d), probably even more pronounced by a presumably overwhelmed health care system having to sustain an even higher peak number (Fig. 6b) of infectives compared to the baseline scenario. However, combining this partial rollback with significantly increased testing activity, isolation of identified cases and reduced contacts with endangered persons (**Scenario RB-IO-IC-T1+**) reduces the expected number of fatalities by a factor of nine (Fig. 6d) and the peak number of known infectives by a factor of two (Fig. 6b), again many more of them only weakly afflicted than in the baseline scenario. Even stricter isolation of cases but less severe restrictions on endangered persons (scenario **RB IC+ IO-T1+**) still results in reductions by a factor of six for fatalities (Fig. 6d) and close to 40% for the maximal number of simultaneously active known infectives (Fig. 6b).

**A close-to-total shutdown of any economic and social activities for three to four months (scenario SD-T) is expected to merely postpone any of the described scenarios**. This is at least the case if we assume that an absolute isolation of Germany as a country will not be possible for a substantial amount of time afterwards, so that the epidemic is bound to inadvertently resurge from endemic reservoirs after the restrictions are being lifted. Even assuming the test activities remain high after rolling back the shutdown, there is no significant effect on either the number of fatalities or the peak number of cases when compared to the baseline scenario with increased testing alone (T1) (results not shown in figures). **The positive effect of a temporary shutdown would be to gain time, which could be an advantage if health systems can be prepared to deal with the disease better, if a vaccine can be developed in time, or if the virus is expected to become less aggressive**. Lacking any information, we did not include these aspects in the model so far.

We should remark that the choice of parameters in our model rests on several assumptions that are not particularly well founded in data. Firstly, **there is significant uncertainty regarding the current number of undetected cases and therefore the current detection ratio**. This clearly affects any assumptions on possible increases of that ratio. In scenarios without additional testing (T0), the total number of infectives at a given time is supposed to be tenfold higher than the number of known cases. Assuming increased testing (T1), the factor reduces to 1.7, while for vastly increased testing (T1+) also of asymptomatic individuals we assume that only 25% of infectives remain undetected, meaning that the number of infectives at a given time is 1.35 times the number of known active cases. The limited capacity of the health care system, in particular of intensive care units, was not yet directly considered as a parameter of our refined model. However, the predicted peak number of infected individuals can be used as a proxy for the expected demand for health care resources at the height of the epidemic. Assuming that a fixed proportion of infected individuals will require intensive care, the maximal number of infectives for a given scenario directly indicates the maximal load on the health care system for this scenario.

The aggressiveness of the virus and hence the mortality among all affected individuals (whether diagnosed or not) is another unknown, but different assumptions about this parameter can be expected to have similar impacts on all the scenarios discussed here.

Given the timing of this study, it is hard to judge the effects of the interventions already in place (in particular CS, HO) on the contact rates with sufficient precision. This uncertainty also affects any predictions about the implications of retracting these measures. **Much will depend on the compliance of the population**. Finally, the effectiveness of case isolation, strongly depending on the detection ratio, is hard to estimate for the above mentioned reasons.

Having said that, **it is obvious from all the simulated scenarios that a combination of vastly increased testing, isolation of known infectives, and restraint in contacts with persons of high age or with relevant preconditions is the most promising approach** if the severity of the epidemic is to be limited to as low a level as possible. **Using antibody tests, as far as available, in addition to direct PCR tests for viral RNA will have the additional benefit of identifying recovered, and hence presumably immune individuals who can then resume their normal levels of activity without jeopardizing any efforts to contain the epidemic**. That even the most optimistic scenarios results in more than ten thousand fatalities and very high numbers of cases is a result of the assumption that the detection ratio is still rather low, albeit increasing, and the presumed number of infectives at the current point in time is already rather large. **Any significant increase in testing is therefore going to reveal a large number of infected individuals already present but still in the dark**. However, the prospect of reducing the number of fatalities by possibly 90% or even more should be a good reason to strongly consider the proposed measures.

Measuring the expected economic and social costs of upholding the current restrictions on movement and contacts and weighing them against the expected gains in terms of decreasing mortality and limiting the excessive demands on the health care system is a very hard challenge that cannot be answered in this study.

For every new unexpected epidemic outbreak long term predictions allow only for rough estimates of the true disease dynamics, hence should be considered carefully. In order to significantly contribute to the control the spread of COVID-19 in Germany, predictions shown in this study will be regularly improved in the coming days and weeks, by enriching the data set, re-calibrating and refining the mathematical model to produce more reliable forecasts.

## Data Availability

The publicly available dataset provided by the Robert Koch Institute (RKI) was used for this study and was last accessed on March 26th, 2020.

https://npgeo-corona-npgeo-de.hub.arcgis.com/datasets/dd4580c810204019a7b8eb3e0b329dd6_0/

## Acknowledgments

The authors would like to thank Prof. Volker Lindenstruth (Frankfurt Institute of Advanced Studies – FIAS) for the constructive discussions initiating this project and his valuable support. We thank Prof. Gordon Pipa of the Institute of Cognitive Sciences at the University of Osnabruück and Dr. Alexander Ullrich of the Robert Koch Institute for providing us with early access to data.

## Contributions

Conceptualization: MVB; Modelling: JF, MVB, JH; Data curation and analysis: NC; Literature research: HVV, JH, JM, NC; Calibration: JF, HVV, JH, MVB, JM. Numerical simulations: JF, JH, HVV, JM, MVB, SK. Writing: MVB, JF, TL.

## Notes

### Competing Interest Statement

The authors have declared no competing interest.

### Funding Statement

no external funding was received

